# “Multimorbidity and consultation time: a systematic review”

**DOI:** 10.1101/19007328

**Authors:** Ana Carolina Reis Tadeu, Inês Rosendo Carvalho e Silva Caetano, Inês Jorge de Figueiredo, Luiz Miguel Santiago

## Abstract

**Background:** Multimorbidity (MM) is one of the major challenges health systems currently face. Management of time length of a medical consultation with a patient with MM is a matter of concern for doctors.

**Objectives:** To describe the impact of MM on the average time of a medical consultation.

**Methods:** A systematic review was performed considering the Preferred Reporting Items for Systematic Review and Meta-analyses (PRISMA) guidelines. The systematic online searches of the Embase and PubMed databases were undertaken, from January 2000 to August 2018. The studies were independently screened by two reviewers to decide which ones met the inclusion criteria. (Kappa=0.84 and Kappa=0.82). Differing opinions were solved by a third person. This systematic review included people with MM criteria as participants (two or more chronic conditions in the same individual). The type of outcome included was explicitly defined – the length of medical appointments with patients with MM. Any strategies aiming to analyse the impact of MM on the average consultation time were considered. The length of time of medical appointment for patients without MM was the comparator criteria. Experimental and observational studies were included.

**Results:** Of 85 articles identified, only 1 observational study was included, showing a clear trend for patients with MM to have longer consultations than patients without MM criteria (*p*<0.001).

**Conclusions:** More studies are necessary to assess better allocation length-time for patients with MM and to measure other characteristics like doctors workload.

## 1. INTRODUCTION

Multimorbidity (MM) is defined by the European General Practice Research Network as “any combination of chronic disease with at least one other disease (acute or chronic) or biopsychosocial factor (associated or not) or somatic risk factor”.^1^ This is sometimes simplified to, “the simultaneous occurrence of two or more chronic diseases in the same individual”.^2^ MM is now one of the main challenges faced by health systems at an international level and occupies a considerable part of the daily activity of General Practitioners/Family Doctors (GPs/FMs) around the world.^3-6^

With an ever ageing world population, MM and its consequences, are becoming a major issue in public health and primary care. According to United Nations data,^7,8^ Europe has the largest percentage of population aged 60 or over (25%).^7^ In 2015 the number of people in the world aged 60 years and older was 901 million.^8^ It is projected that in 2030 this figure will rise to 1.4 billion (a 56% increase since 2015) and stand at 2.1 billion in 2050.^8^ Several studies have shown that there is a significant association between age^2^ and the prevalence of MM, most national health systems not being prepared or able to cope with this rapid ageing with many demands.^5,6^

So it is imperative to think about the approach to patients with MM to maximize the quality of services provided by the Health Services (HS) consequently guaranteeing a better quality of life for such patients.

GPs/FMs medical team face various difficulties in caring for a patient with MM like lack of resources; consultation time restrictions; lack of interdisciplinary care/teams; inadequate patient support (largely relying on community-based support services); inadequate tools (guidelines are drawn up strictly for specific diseases and not for the MM patient); the attitude of the patient (often discouraged and poorly engaged).^4,9^

Information about the length-time of a consultation with a patient with MM is essential to better organize and deliver healthcare. To our knowledge, no previous review has summarized data related to: “What is the impact of having MM on the medical consultation?” “Is the average length-time consultation with a patient with MM longer than for a patient without MM?”

## 2. METHODS

A systematic review was performed considering the Preferred Reporting Items for Systematic Review and Meta-analyses (PRISMA) guidelines for systematic reviews and meta-analysis with:

### 2.1 Eligibility criteria

Articles about people with MM. The most widely used definition of MM was used, which is the coexistence of two or more chronic conditions in the same individual.^2^ The World Health Organization (WHO) definition of chronic disease was adopted, namely, “health problems that require ongoing management over a period of years or decades”.^10^

Studied outcome: explicitly defined length-time of medical appointments with patients with MM. Any strategies aiming to analyse the impact of MM on the average consultation time were considered. Studies which did not specify the time spent on medical appointments were excluded.

The length of time of medical appointment for patients without MM was the comparator criteria.

Experimental and observational studies were included.

### 2.2 Information sources and search strategy

The systematic online searches were undertaken using combinations of keywords in the following electronic databases: the Embase and PubMed databases, from 1^st^ January 2000 until the 31^st^ August 2018 to find pertinent studies.

The search was limited to papers in English, Portuguese, Spanish and French. No other limits were imposed during this stage of the study.

### 2.3 Data extraction and quality assessment

The potentially relevant studies were selected in two stages. First, the titles and abstracts quoted in the literature search were independently screened by two reviewers (CT and IF) to decide which ones met the inclusion criteria (Kappa=0.84). Those not meeting the inclusion criteria were excluded. Differing opinions on studies inclusion were resolved by a third person (IR).

Secondly, the researchers independently read and analysed the integrity of the matching studies and tried to reach an agreement concerning eligibility (Kappa=0.82). Those not meeting the inclusion criteria were excluded. The quality and risk of bias of the included studies was assessed using the Newcastle-Ottawa Scale (NOS), more precisely, the Newcastle-Ottawa scale adapted for cross-sectional studies.^11^ This tool assesses three aspects of a study: the selection of the sample; the comparability of the groups; and the outcome (assessment of outcome and statistical test). It is composed of 7 items and classifies the study in 4 possible levels: Very good (9-10 points), Good (7-8 points), Satisfactory (5-6 points) and Unsatisfactory (0-4 points). Any disagreement was resolved through consensus.

This systematic review was conducted using Covidence13, the standard production platform used for Cochrane reviews, which was used for the data and records management.

### 2.4 Outcomes and statistical analysis

The patients were split into two groups, those with and those without MM, and the relative frequencies were calculated. The results were analyzed using the chi-square distribution test.

## 3. RESULTS

### 3.1 Study selection

As described in **Flowchart 1**, the electronic database searches started with 85 potentially eligible references (26 in PubMed and 59 in Embase). Of these, 5 were duplicates and were thus excluded and 31 were considered irrelevant based on a review of the title and abstract. The remnant studies were fully read, analysed and assessed for eligibility, 36 being excluded due to wrong outcome^3,12-46^, 5 because of wrong study design^47-51^, 4 due to wrong patient population^9,52-54^, 2 for wrong language^55-56^ and 1 for wrong setting^57^. In the end, 1 study was included.^58^

**Flowchart 1.**
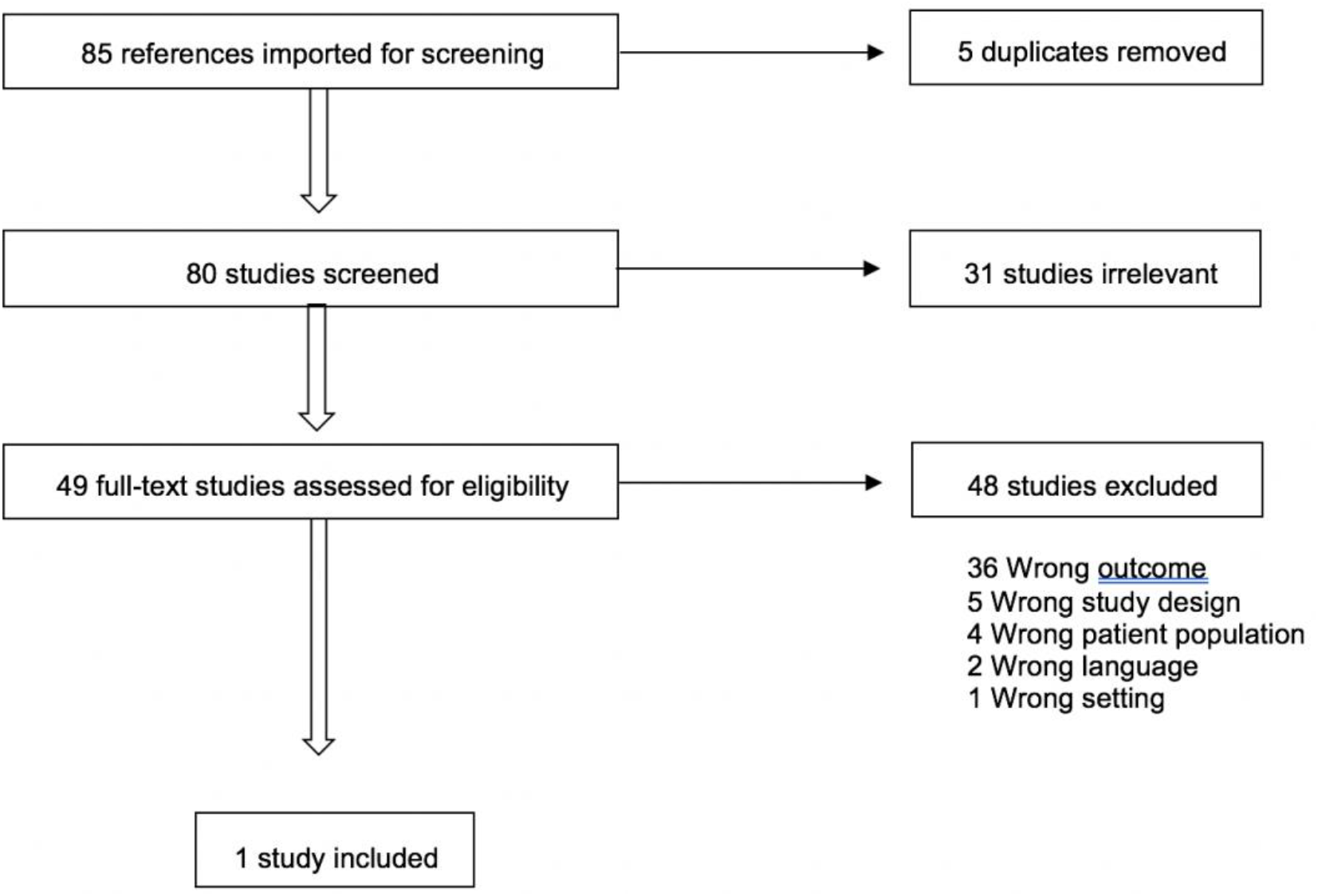
Literature search and selection process for studies included.

### 3.2 Study characteristics and quality

The main relevant features and outputs of the study were extracted for the purpose of this systematic review and are summed up in **Table 1**.

**Table 1.** Summary of study’s characteristics.

| Author | Year of study | Country | Design | Number of participants | Population (inclusion criteria) | Setting | Method of data collection | Outcomes measured | Author's conclusions |
| --- | --- | --- | --- | --- | --- | --- | --- | --- | --- |
| Moth et al | 2008-2009 | Denmark | Cross-sectional | 404 GPs, 8236 contacts | Persons aged 40 years or more | General practice | Registration form completed by GP about all patient contacts on one randomly assigned date during the study period. | - Length of consultation time, chronic disease, reason for appointment, diagnosis, number of additional psychosocial problems raised by the patient during the consultation, difficulty found with consultation of the consultation, referral to specialized care, and whether a nurse could have replaced the GP. | - GPs found consultations with patients suffering from chronic conditions to be more difficult than those with patients without chronic disease ($p < 0.001$). |

GP – General Practitioner.

The included study was conducted between 2008 and 2009 in Denmark, over 12 months. It involved 404 general practitioners (GPs) participants and a total of 8236 contacts. It included patients aged 40 years or more, grouped as those without any chronic condition and those with one, two, three or more chronic conditions.

During the study period, the GPs completed a one-page registration form for each of their patient contacts. Of the various items that were registered, the ones relevant forthis review were information on chronic diseases and length-time of consultation. Quality assessment result, performed as described in methodology, is presented in **Table 2**. The quality of the study was considered satisfactory (score 6 out of a maximum score of 10), its main weakness being in the comparability section.

**Table 2.** Quality of study - Newcastle-Ottawa Scale adapted for cross-sectional studies.

| Author | Selection |  |  |  | Comparability | Outcome |  | Total score | Power |
| --- | --- | --- | --- | --- | --- | --- | --- | --- | --- |
|  | Representativeness of the sample | Sample size | Non-respondents | Ascertainment of exposure (risk factor) | Comparability of subjects in different outcome groups on the basis of design or analysis. | Assessment of outcome | Statistical test |  |  |
| Moth et al | * | - (b) | * | ** | - | * | * | 6* | Satisfactory |

(b) – Calculation not reported.

### 3.3 Results of study

**Table 3** shows the relationship between the length of consultation time and the type of patient (with and without MM). There is a clear significant trend for patients with MM to have longer consultations than patients without MM (*p<*0.001).

| Length of consultation time | No MM |  | MM |  | *p*-value |
| --- | --- | --- | --- | --- | --- |
|  | (n) | % | (n) | % |  |
| <5 min | 293 | 11.7 | 96 | 7.7 | $p<0.001$ |
| 6-15 min | 1686 | 67.3 | 804 | 64.9 |  |
| 16-30 min | 485 | 19.4 | 314 | 25.3 |  |
| >30 min | 42 | 1.7 | 25 | 2.0 |  |
| Total | 2506 | 100 | 1239 | 100 |  |

More than 25% of MM patients had a consultation length-time of at least 16 minutes while more than 75% of the patients without MM had a consultation length-time under 15 minutes. Length-time consultation on both types of patients is more frequently between 6 and 15 minutes. There is a significant difference, however, in the percentage of patients with MM requiring more time than patients without MM criteria.

## 4. DISCUSSION

The present systematic review sought to answer the following question: “Is the average consultation time spent on patients with MM longer than that spent on patients who do not meet the MM criteria?” could only identify one study,^58^ undertaken in Denmark, in which the consultation time was logged as a function of the number of chronic diseases. This study revealed a tendency for consultations to take longer for patients with MM than for those without MM. However, the study was not directly aimed at answering this question and it did not take confounding factors into account. In addition, it does not describe the calculation to determine the sample size of the study and it could be inaccurate to study this specific outcome. So the study was classified as “Satisfactory”.

The small number of publications in the literature on this subject shows that more studies should be designed to investigate the impact of MM on the consultation length-time. It is vital to analyse this issue in order to manage resources so that they meet the actual need, and to ensure the services provided by health services, national or private, are appropriate. It will thus be possible to guarantee better quality health services and outcomes for these patients. In fact MM is about a patient with more then two chronic diseases or one chronic disease with biopsychosocial factor (associated or not) or somatic risk factor, so illness being included and not only about the specific sufferances.^1,9^

In studies with calculated size population representative random samples it is important that accurate methods are used to measure the real length-time of a consultation (i.e. from the moment that the doctor opens the patient’s file to the moment it is closed). Using stop-watches and self-reporting will probably lead to inaccuracies. The calculation of the length-time of the consultation obtained by dividing the total time a medical practitioner is in the clinic by the number of patients could yield average times that mask the real duration of each patient’s consultation. Also confounding factors must be eliminated as the time spent on administrative work, breaks and work meetings. Only direct observation using video recording has been proven to obtain accurate values when measuring the duration of consultations,^59^ which could be a procedure that mitigates many of the errors previously mentioned. The length-time of a consultation must be measured accurately to avoid errors and skewed judgements. It is essential to identify beforehand any possible confounding factors inherent to the patients (for example, hearing difficulty, education level, age, socio-economic level), inherent to the doctor (in particular, a change in behaviour due to the participation in the research study – Hawthorne effect^60^), and inherent to the consultation/institution (for example, glitches in computer systems, organization of necessary information in the health informatics records, coding errors, telephone call interruptions). The time lost searching for information in consultation, the friendliness of clinical informatics and the time spent on e.records are also issues to be studied and thought of ^61, 62^. Health determinants are factors to be studied in such a MM population for better health and social outcomes.^63^

The data analysis must be evaluated using objective validated laboratory methods and, if possible, it should be a blind assessment. Statistical tests used to analyse the data must be appropriate and clearly described. Measures of association, including confidence intervals and the P value, must be presented.

The main limitation of this systematic review was the difficulty in ensuring that all the relevant literature was included. Even though the research used two of the main databases – Pubmed and Embase – there could be other relevant material in grey literature.

The scarcity of the literature that was found was a limitation for this review. The one publication found, besides not directly answering our question, also does not take confounding factors into account, and does not describe the calculation to determine the sample size of the study. However, it does highlight the relevance of the subject matter.

## 5. CONCLUSIONS

This “impact of MM on the duration of a consultation” has hardly been studied, this systematic review shows.

A tendency for consultations of patients with MM to take longer than those without MM was found in the only one study with “satisfactory” quality which met the inclusion criteria.

So more research is needed to acquire more information on this subject, important to deal not only with diseases but with the person suffering from MM for consultation must have the adequate length duration to enable doctors and stake-holders with a proper quantification of the time and associated costs.

If a longer consultation time is confirmed, it will be important to rethink and adapt GPs’ lists of patients in order to achieve better medical care providing agendas with specific times and allocating enough time for all the required tasks.

## Data Availability

All data will be available upon request

## 6. LIST OF ABBREVIATIONS

MM: Multimorbidity
GP: General Practitioner
PRISMA: Preferred Reporting Items for Systematic Review and Meta-analyses
HS: Health Service
WHO: World Health Organization

## 7. DECLARATIONS

Ethics approval and consent to participate – Not applicable

Consent for publication – Not applicable

Availability of data and materials – Materials can be assessed upon recquest

Financial and non-financial competing interests

No author states financial and non-financial competing interests for this work.

## Authors’ contributions

ACRT: Data extraction, data examination, treatment of data, writting of manuscript and its scientific ctiticism as well as aproval.

IRCSC: Data extraction, data examination, treatment of data, writting of manuscript and its scientific ctiticism as well as aproval.

Inês Jorge de Figueiredo: Sata examination, treatment of data, writting of manuscript and its scientific ctiticism as well as aproval.

Luiz Miguel Santiago: Scientific review of the article.

All authors read and approved the final manuscript.

### Acknowledgements

Helena Donato, MD for her valuable contribution in the literature search.

## Group authorship

Not aplicable

